# Mapping Health-Related Quality of Life in Mississippi: Longitudinal Spatial Clustering and Socioeconomic Drivers

**DOI:** 10.64898/2026.01.31.26345277

**Authors:** Jae Eun Lee, JungHye Sung, Ji-Young Lee

**Author notes:** Author Contributions Dr. Jae Eun Lee conceived and designed the study, performed all spatial analyses (including GIS mapping, LISA clustering, choropleth mapping, and spatial lag panel regression), conducted the primary modeling, created all figures and tables, and drafted the manuscript. Dr. Junghye Sung acquired and harmonized the data, performed the statistical analyses, interpreted the results, and drafted and critically revised the manuscript for important intellectual content. Dr. Ji-Young Lee interpreted the results, helped draft the manuscript and critically revised it for important intellectual content. Drs. Barner, Offiah, and Hays helped draft the manuscript and critically revised it for important intellectual content. All authors reviewed and approved the final version of the manuscript and agree to be accountable for all aspects of the work.

## Abstract

**Background:** Mississippi consistently ranks among the lowest U.S. states in health-related quality of life (HRQoL) outcomes, with disparities most severe in rural and socioeconomically disadvantaged counties, particularly the Mississippi Delta. Understanding longitudinal spatial and temporal patterns is essential for equity-focused public health strategies and chronic disease prevention.

**Objective:** This study examined geographic disparities in HRQoL across Mississippi’s 82 counties from 2015 to 2025 to identify persistent hotspots, assess convergence with national averages, and inform targeted interventions.

**Methods:** County-level data from the County Health Rankings & Roadmaps (2015–2025 releases) were used to construct an annual principal component analysis–derived composite HRQoL score from Behavioral Risk Factor Surveillance System measures (percentage reporting poor or fair health, physically unhealthy days, and mentally unhealthy days). Spatial patterns were assessed using choropleth maps, Moran’s I, and Local Indicators of Spatial Association (LISA). A spatial autoregressive lag model with year fixed effects identified key drivers.

**Results:** Mississippi showed persistent disadvantage in poor or fair health (stable gap of 0.06–0.07 percentage points above national averages) but recent convergence in physically unhealthy days and reversal in mentally unhealthy days (fewer days in disadvantaged subgroups, e.g., high child poverty and low education). LISA maps revealed enduring High-High hotspots of poor HRQoL in the Delta region, while Low-Low cold spots along the Gulf Coast contracted substantially by 2025 (many formerly advantaged counties now non-significant). The spatial lag model confirmed significant dependence (ρ = 0.13, P < .001), with adult smoking and uninsurance as leading modifiable predictors.

**Conclusions:** Mississippi’s HRQoL trajectory shows symptom-based improvements alongside enduring structural disparities in self-rated health and persistent geographic inequities. Expanding smoke-free policies (particularly in the Delta), strengthening primary care access, and increasing insurance coverage represent high-impact strategies to reduce disparities, advance health equity, and support chronic disease prevention in high-burden regions of the U.S. South.

## Introduction

Health-related quality of life (HRQoL) is a multidimensional construct encompassing physical, mental, and social well-being, often assessed via self-reported measures such as the CDC’s Healthy Days from the Behavioral Risk Factor Surveillance System (BRFSS).

HRQoL is strongly influenced by socioeconomic factors, including income, education, rural residence, and access to healthcare, which contribute to elevated chronic disease risks and poorer outcomes in underserved regions (1).

Mississippi consistently ranks among the lowest U.S. states in HRQoL and related health outcomes, with severe disparities in rural and socioeconomically disadvantaged counties, particularly the Mississippi Delta. The state has historically reported higher rates of poor physical and mental unhealthy days, fair/poor self-rated health, and chronic conditions like diabetes and smoking-related diseases compared to national averages (2, 3). These gaps are driven by structural inequities, limited healthcare access, and social determinants of health (SDOH), with life expectancy in some rural areas lagging several years behind urban U.S. norms.

Despite growing HRQoL research, significant gaps persist in longitudinal spatial analyses of multidimensional HRQoL patterns, especially in high-disparity states like Mississippi. Nationwide studies have identified low-HRQoL clusters in the Southeast, including the Delta, but applications of advanced clustering (e.g., LISA) and modeling to integrated BRFSS-derived data from the County Health Rankings & Roadmaps (CHR&R) remain scarce (4, 5). Most work emphasizes aggregate or disease-specific outcomes rather than broad, temporal trends of composite HRQoL, leaving policymakers without targeted insights for equity-focused interventions amid recent post-pandemic shifts.

This novel study addresses these critical gaps by examining longitudinal trends and geographic disparities in HRQoL across Mississippi’s 82 counties from 2015 to 2025. Using a PCA-derived composite HRQoL index and advanced spatial analyses, it identifies persistent hotspots, evaluates convergence with national averages, and uncovers key drivers to inform precise, equity-focused public health strategies and chronic disease prevention in high-burden regions.

## Methods

### Study Setting and Data Source

This study utilized county-level data for Mississippi from the County Health Rankings & Roadmaps (CHR&R) program for release years 2015 through 2025 (6). The CHR&R dataset, produced annually by the Robert Wood Johnson Foundation in partnership with the University of Wisconsin Population Health Institute, provided standardized measures derived from national surveys and administrative sources, including the Behavioral Risk Factor Surveillance System (BRFSS) (7), National Center for Health Statistics mortality files, and U.S. Census Bureau data (8). Analyses were restricted to Mississippi’s 82 counties. Data were accessed in December 2025. As the data were publicly available and de-identified, institutional review board approval was not required.

Health-Related Quality of Life Measures

Three core HRQoL measures from the CHR&R dataset were used (6):

- Percentage of adults reporting fair or poor health (self-rated overall health)
- Average number of physically unhealthy days reported in the past 30 days
- Average number of mentally unhealthy days reported in the past 30 days

These indicators were derived from the Centers for Disease Control and Prevention’s BRFSS survey and employed small-area estimation techniques to produce reliable county-level estimates (7). Higher values indicated poorer HRQoL. Due to multi-year aggregation and processing, CHR&R measures incorporated a 2–3 year lag (e.g., the 2025 release primarily reflected conditions circa 2022).

### Composite HRQoL Index

A multidimensional HRQoL measure was constructed by applying principal component analysis (PCA) to the three indicators: percentage of adults reporting fair or poor health, average physically unhealthy days, and average mentally unhealthy days. Each variable was standardized to z-scores within its release year for comparability, and PCA was conducted separately for each year from 2015 through 2025. The first principal component (PC1) was retained as the composite score (hrqol_pca), with higher values indicating worse HRQoL. PC1 explained a large proportion of the variance across years (59.2%–97.4%, mean 86.2%), confirming its robustness as a summary measure. PCA was selected over simple summation or arbitrary weighting because it was data-driven, maximized shared variance among correlated indicators, and preserved temporal variation without subjective weights. The resulting hrqol_pca scores were merged back into the Mississippi dataset by county and release year. The composite demonstrated strong construct validity, with Pearson correlations of r = 0.90 with poor or fair health, r = 0.77 with physically unhealthy days, and r = 0.46 with mentally unhealthy days (pooled across all county-years; all p < 0.001).

### Covariates

Covariates were selected to represent key domains from the CHR&R framework: health behaviors, clinical care, and social/economic factors (8). These included ratio of population to dentists, preventable hospital stays, adult smoking rate, diabetes prevalence, injury deaths (per 100,000), percentage non-Hispanic Black, percentage aged 65 and older, uninsured adults (percentage), and median household income. Missing covariate values were imputed using the overall median within each release year.

### Subgroup Analyses

To examine whether recent HRQoL convergence varied by socioeconomic context, 11 key socioeconomic indicators (e.g., uninsured children/adults, unemployment, educational attainment, median household income, child poverty, racial composition, age distribution) were dichotomized annually within each release year. Cutpoints were set at the median of non-missing values separately for Mississippi counties and all non-Mississippi U.S. counties, creating comparable “high” and “low” strata that accounted for temporal and distributional differences.

Pooled means (with standard deviations) for the three core HRQoL measures (poor or fair health percentage, physically unhealthy days, mentally unhealthy days) were calculated across the most recent three releases (2023–2025, reflecting ∼2020–2022 conditions) for each subgroup. Mean differences (gaps: Mississippi minus non-Mississippi) were computed, along with statistical significance (p-values) and effect sizes (Cohen’s d) to quantify the magnitude of disparities or convergence/reversal between Mississippi and the rest of the U.S. within high- and low-burden strata.

### Spatial Analyses

County boundaries were obtained from the 2023 U.S. Census TIGER/Line shapefiles (9). Contiguity-based Queen spatial weights (row-standardized) were constructed using point centroids for consistency across years (10). Global spatial autocorrelation was assessed annually using Moran’s I (11). Local Indicators of Spatial Association (LISA) were computed with 999 permutations and a significance threshold of p < 0.05 (10). Choropleth maps visualized the composite HRQoL score, with a common scale applied across selected years for comparability.

A spatial autoregressive lag (SAR lag) model with year fixed effects (2015 as reference) was estimated via maximum likelihood estimation, using the composite HRQoL score as the dependent variable (12, 13). The model specification included the selected covariates and a spatially lagged dependent variable to account for spillover effects from neighboring counties, addressing spatial dependence observed in Moran’s I and LISA results. Year fixed effects controlled for temporal trends across the panel. Multicollinearity was assessed using variance inflation factors (VIF < 5 for all covariates), and coefficients represented direct effects on HRQoL (higher values indicating worse outcomes). The approach was selected over spatial error or other specifications based on its suitability for capturing substantive spatial spillovers in health outcomes.

### Statistical Software

All analyses were conducted in Python 3.11. Data management and preprocessing used pandas, numpy, and geopandas. PCA was performed with scikit-learn. Spatial weights, Moran’s I, LISA, and SAR lag modeling used libpysal, esda, and spreg. Choropleth maps were generated with matplotlib and geopandas.

## Results

### Trends in Health-Related Quality of Life Measures

Table 1 displays annual averages for the three core health-related quality of life (HRQoL) measures in Mississippi compared to U.S. national averages from 2015 to 2025 releases, along with the gap (Mississippi minus U.S.; positive values indicate worse outcomes in Mississippi). Mississippi consistently reported higher average poor physical health days than the national average in most releases, with the gap ranging from 0.00 (2023 release, reflecting ∼2020 conditions) to 0.81 days (2020 release, reflecting ∼2017 conditions). The largest gaps appeared in releases corresponding to the COVID-19 period (2020–2021 releases), but disparities narrowed substantially in more recent releases.

**Table 1.** Trends in Health-Related Quality of Life Measures: Mississippi vs. U.S. Averages, 2015–2025.

| Year | Poor Physical Health Days |  |  | Poor Mental Health Days |  |  | Poor or Fair Health (%) |  |  |
| --- | --- | --- | --- | --- | --- | --- | --- | --- | --- |
|  | MS | Non-MS | Gap | MS | Non-MS | Gap | MS | Non-MS | Gap |
| 2015 | 4.37 | 3.81 | 0.56 | 4.15 | 3.54 | 0.60 | 0.23 | 0.17 | 0.06 |
| 2016 | 4.26 | 3.80 | 0.46 | 4.09 | 3.66 | 0.43 | 0.23 | 0.17 | 0.06 |
| 2017 | 4.57 | 3.90 | 0.67 | 4.32 | 3.77 | 0.55 | 0.24 | 0.17 | 0.07 |
| 2018 | 4.32 | 3.91 | 0.41 | 4.26 | 3.92 | 0.34 | 0.24 | 0.17 | 0.06 |
| 2019 | 4.32 | 3.91 | 0.41 | 4.26 | 3.92 | 0.34 | 0.24 | 0.17 | 0.06 |
| 2020 | 4.78 | 3.97 | 0.81 | 4.72 | 4.15 | 0.57 | 0.25 | 0.18 | 0.07 |
| 2021 | 5.01 | 4.37 | 0.63 | 5.25 | 4.66 | 0.59 | 0.27 | 0.20 | 0.07 |
| 2022 | 4.70 | 4.33 | 0.37 | 5.50 | 4.87 | 0.63 | 0.27 | 0.20 | 0.06 |
| 2023 | 3.52 | 3.52 | 0.00 | 4.56 | 4.81 | -0.25 | 0.22 | 0.16 | 0.06 |
| 2024 | 4.15 | 3.89 | 0.26 | 5.04 | 5.22 | -0.18 | 0.25 | 0.18 | 0.07 |
| 2025 | 4.48 | 4.46 | 0.02 | 5.37 | 5.65 | -0.28 | 0.25 | 0.19 | 0.06 |

Note that these measures are derived from Behavioral Risk Factor Surveillance System (BRFSS) data and typically reflect conditions with a 2–3 year lag (e.g., the 2025 release primarily uses 2022 data, while earlier releases aggregate multiple prior years).

For poor mental health days, Mississippi also showed higher values in earlier releases (gap 0.34–0.63 days), but from the 2023 release onward (reflecting ∼2020–2022 conditions), the state reported fewer poor mental health days than the U.S. average (negative gap of -0.18 to -0.28 days), suggesting relative improvement in recent years. The percentage of adults reporting poor or fair health remained persistently higher in Mississippi across all releases, with a stable gap of approximately 0.06–0.07 percentage points, indicating a chronic disparity in self-rated overall health that shows little temporal variation.

These trends highlight ongoing HRQoL challenges in Mississippi—particularly in self-rated health—alongside evidence of convergence with national averages in physical and mental health days in releases reflecting post-2020 conditions.

### Subgroup Variation in Recent Convergence of HRQoL Measures

Table 2 presents pooled mean values for the three HRQoL measures across the most recent three releases (2023–2025, reflecting approximately 2020–2022 conditions), comparing Mississippi counties to all non-Mississippi U.S. counties within high vs. low strata of key socioeconomic and demographic characteristics (mean differences, p-values, and Cohen’s d reported).

**Table 2.** Subgroup Differences in Health-Related Quality of Life Measures Between Mississippi and Non-Mississippi U.S. Counties (Pooled 2023–2025 Releases)

| Subgroup | Poor Physical Health Days |  |  |  | Poor Mental Health Days |  |  |  | Quality of Life |  |  |  |
| --- | --- | --- | --- | --- | --- | --- | --- | --- | --- | --- | --- | --- |
|  | Non-MS | MS | Gap | Cohen's d | Non-MS | MS | Gap | Cohen's d | Non-MS | MS | Gap | Cohen's d |
|  | Mean(sd) | Mean(sd) | Mean(sd) |  | Mean(sd) | Mean(sd) | Mean(sd) |  | Mean(sd) | Mean(sd) | Mean(sd) |  |
| Uninsured Children – Low | 3.88 (0.80) | 4.04 (0.59) | +0.16 (0.05)** | 0.20 | 5.25 (0.73) | 4.95 (0.45) | -0.30 (0.04)*** | 0.41 | 0.17 (0.05) | 0.24 (0.05) | +0.07 (0.00)*** | 1.58 |
| Uninsured Children – High | 4.03 (0.72) | 4.06 (0.51) | +0.03 (0.05) | 0.04 | 5.20 (0.69) | 5.03 (0.37) | -0.18 (0.04)*** | 0.26 | 0.18 (0.05) | 0.23 (0.04) | +0.05 (0.00)*** | 1.13 |
| Uninsured Adults – Low | 3.76 (0.77) | 3.87 (0.57) | +0.11 (0.05)* | 0.14 | 5.13 (0.73) | 4.90 (0.43) | -0.23 (0.04)*** | 0.32 | 0.16 (0.04) | 0.23 (0.05) | +0.07 (0.00)*** | 1.52 |
| Uninsured Adults – High | 4.15 (0.70) | 4.23 (0.46) | +0.08 (0.04) | 0.11 | 5.32 (0.67) | 5.08 (0.38) | -0.24 (0.04)*** | 0.36 | 0.19 (0.05) | 0.25 (0.04) | +0.06 (0.00)*** | 1.36 |
| Unemployment – Low | 3.72 (0.68) | 3.85 (0.53) | +0.13 (0.05)** | 0.19 | 5.01 (0.68) | 4.93 (0.40) | -0.08 (0.04)* | 0.13 | 0.16 (0.04) | 0.22 (0.03) | +0.06 (0.00)*** | 1.39 |
| Unemployment – High | 4.20 (0.77) | 4.25 (0.50) | +0.05 (0.05) | 0.07 | 5.44 (0.68) | 5.05 (0.42) | -0.39 (0.04)*** | 0.59 | 0.19 (0.05) | 0.26 (0.04) | +0.07 (0.00)*** | 1.48 |
| Some College – Low | 4.37 (0.68) | 4.26 (0.48) | -0.11 (0.04)* | 0.17 | 5.53 (0.65) | 5.05 (0.41) | -0.48 (0.04)*** | 0.75 | 0.20 (0.04) | 0.26 (0.04) | +0.06 (0.00)*** | 1.33 |
| Some College – High | 3.54 (0.60) | 3.84 (0.53) | +0.30 (0.05)*** | 0.51 | 4.92 (0.64) | 4.92 (0.41) | +0.00 (0.04) | 0.01 | 0.15 (0.03) | 0.22 (0.04) | +0.07 (0.00)*** | 2.21 |
| Median Household Income – Low | 4.35 (0.71) | 4.28 (0.48) | -0.07 (0.04) | 0.10 | 5.51 (0.68) | 5.04 (0.42) | -0.47 (0.04)*** | 0.70 | 0.20 (0.04) | 0.27 (0.04) | +0.06 (0.00)*** | 1.45 |
| Median Household Income – High | 3.56 (0.59) | 3.82 (0.52) | +0.25 (0.05)*** | 0.43 | 4.94 (0.62) | 4.94 (0.41) | -0.00 (0.04) | 0.01 | 0.15 (0.03) | 0.21 (0.03) | +0.06 (0.00)*** | 1.94 |
| High School – Low | 3.91 (0.75) | 4.16 (0.50) | +0.25 (0.04)*** | 0.34 | 5.15 (0.72) | 5.02 (0.40) | -0.13 (0.04)*** | 0.18 | 0.17 (0.05) | 0.25 (0.04) | +0.08 (0.00)*** | 1.69 |
| High School – High | 4.03 (0.79) | 3.92 (0.57) | -0.12 (0.06)* | 0.15 | 5.35 (0.68) | 4.95 (0.44) | -0.41 (0.04)*** | 0.60 | 0.18 (0.05) | 0.22 (0.04) | +0.04 (0.00)*** | 0.94 |
| Children Poverty – Low | 3.53 (0.58) | 3.85 (0.53) | +0.31 (0.05)*** | 0.53 | 4.90 (0.62) | 4.95 (0.42) | +0.05 (0.04) | 0.08 | 0.15 (0.03) | 0.21 (0.03) | +0.07 (0.00)*** | 2.20 |
| Children Poverty – High | 4.38 (0.68) | 4.26 (0.49) | -0.12 (0.05)** | 0.18 | 5.56 (0.64) | 5.03 (0.41) | -0.53 (0.04)*** | 0.83 | 0.21 (0.04) | 0.27 (0.04) | +0.06 (0.00)*** | 1.45 |
| % Non-Hispanic White – Low | 4.04 (0.73) | 4.19 (0.55) | +0.15 (0.05)** | 0.20 | 5.28 (0.65) | 4.98 (0.43) | -0.30 (0.04)*** | 0.47 | 0.19 (0.05) | 0.26 (0.04) | +0.07 (0.00)*** | 1.53 |
| % Non-Hispanic White – High | 3.87 (0.78) | 3.91 (0.52) | +0.04 (0.05) | 0.05 | 5.18 (0.77) | 5.00 (0.40) | -0.17 (0.04)*** | 0.23 | 0.16 (0.04) | 0.22 (0.03) | +0.05 (0.00)*** | 1.26 |
| % Non-Hispanic Black – Low | 3.86 (0.79) | 3.89 (0.51) | +0.03 (0.05) | 0.04 | 5.11 (0.75) | 4.99 (0.40) | -0.12 (0.04)** | 0.16 | 0.17 (0.05) | 0.21 (0.03) | +0.05 (0.00)*** | 1.05 |
| % Non-Hispanic Black – High | 4.05 (0.73) | 4.21 (0.54) | +0.16 (0.05)** | 0.22 | 5.34 (0.65) | 4.99 (0.43) | -0.36 (0.04)*** | 0.55 | 0.19 (0.05) | 0.26 (0.04) | +0.08 (0.00)*** | 1.71 |
| % Below 18 Years Of Age – Low | 3.93 (0.74) | 4.04 (0.50) | +0.11 (0.05)* | 0.14 | 5.27 (0.69) | 4.96 (0.40) | -0.32 (0.04)*** | 0.46 | 0.17 (0.04) | 0.24 (0.04) | +0.07 (0.00)*** | 1.47 |
| % Below 18 Years Of Age – High | 3.98 (0.78) | 4.06 (0.59) | +0.08 (0.05) | 0.10 | 5.18 (0.73) | 5.02 (0.43) | -0.16 (0.04)*** | 0.22 | 0.18 (0.05) | 0.24 (0.05) | +0.06 (0.00)*** | 1.26 |
| % 65 And Older – Low | 3.99 (0.78) | 3.98 (0.60) | -0.01 (0.05) | 0.01 | 5.25 (0.68) | 4.95 (0.42) | -0.30 (0.04)*** | 0.44 | 0.18 (0.05) | 0.23 (0.05) | +0.05 (0.00)*** | 1.07 |
| % 65 And Older – High | 3.92 (0.75) | 4.12 (0.49) | +0.20 (0.05)*** | 0.27 | 5.21 (0.74) | 5.02 (0.40) | -0.18 (0.04)*** | 0.25 | 0.17 (0.04) | 0.24 (0.04) | +0.07 (0.00)*** | 1.68 |

Note. a Data are pooled means (SD) from County Health Rankings & Roadmaps releases 2023–2025 (reflecting approximately 2020–2022 conditions). Gaps = Mississippi minus non-Mississippi U.S. counties. b Significance: \*P < .05, \*\*P < .01, \*\*\*P < .001 (t-test for mean differences). c Cohen's d effect size: values >0.5 indicate moderate to large effects. d Abbreviations: Non-MS = non-Mississippi U.S. counties; MS = Mississippi counties; SD = standard deviation

Poor or fair health exhibited the largest and most consistent disparities, with Mississippi counties reporting significantly higher percentages across all high-burden subgroups (Cohen’s d = 1.07–2.21). The gap was particularly large in relatively advantaged subgroups (high educational attainment, low child poverty, higher median household income), indicating that long-term structural disadvantages continue to impair perceived overall health despite some socioeconomic strengths.

Poor mental health days showed a striking reversal: in nearly every high-burden subgroup, Mississippi reported fewer days than comparable non-Mississippi counties (moderate to large effect sizes, d = 0.23–0.83). The largest reversals occurred in the most disadvantaged communities (high child poverty, d = 0.83; low educational attainment, d = 0.75; low median household income, d = 0.70; high unemployment, d = 0.59; high % non-Hispanic Black, d = 0.55), suggesting better recent mental well-being conditional on socioeconomic disadvantage.

Poor physical health days displayed mixed convergence. The statewide gap narrowed, with many high-burden subgroups showing small or non-significant differences (e.g., high uninsurance, high unemployment, high % Non-Hispanic White). Several subgroups even favored Mississippi (negative gaps), particularly high child poverty and low education/income strata. However, larger positive gaps persisted in advantaged subgroups (e.g., low child poverty, d = 0.53; high some college, d = 0.51), indicating that convergence was driven primarily by improvements in Mississippi’s most disadvantaged communities.

### Geographic Distribution of Composite HRQoL (2016 and 2025)

Figure 1 present choropleth maps of the PCA-based composite HRQoL score for Mississippi counties in 2016 (left panel) and 2025 (right panel), with higher scores (redder shades) indicating worse health-related quality of life and lower scores (bluer shades) indicating better HRQoL.

**Figure 1.**
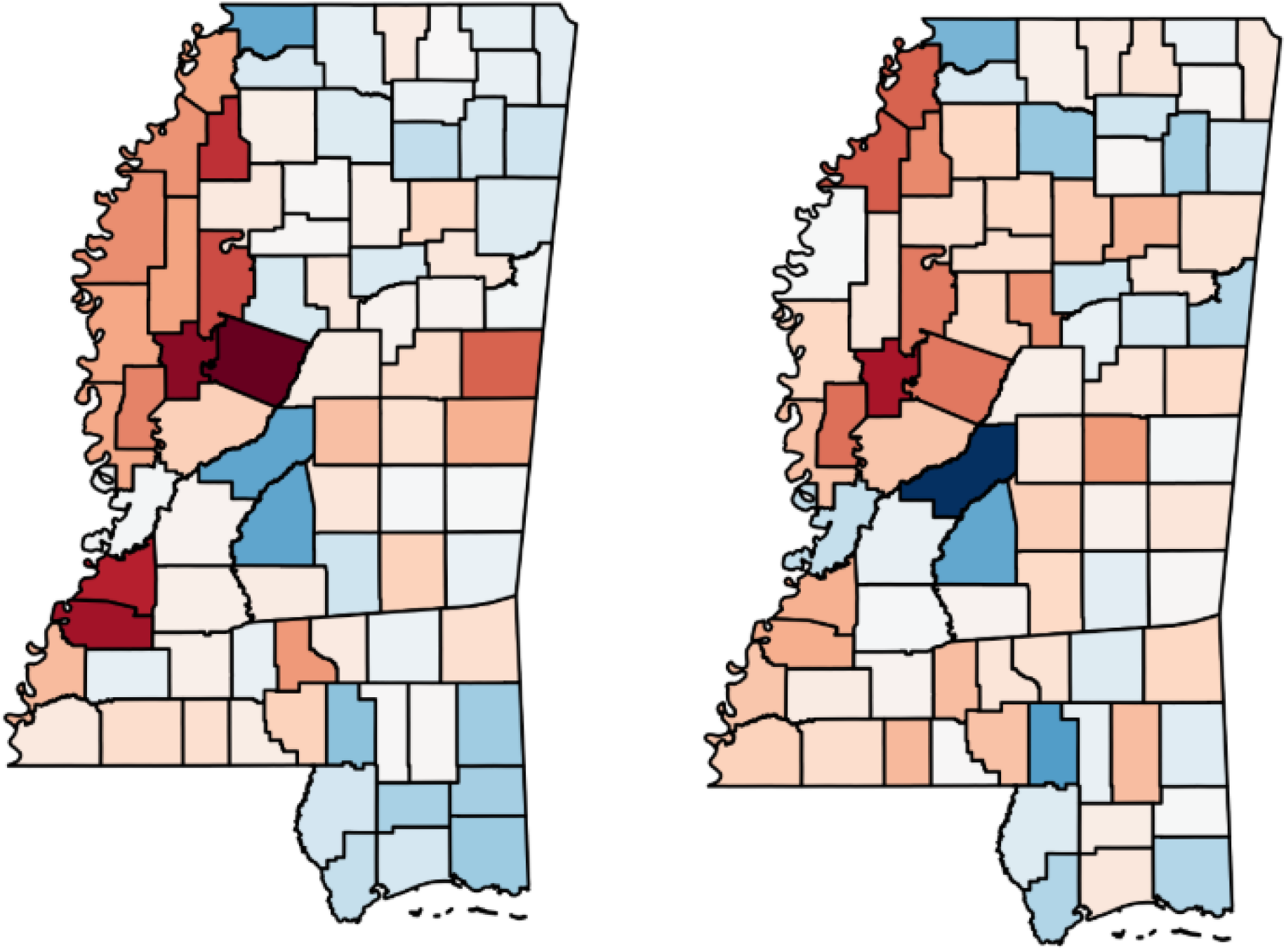
Choropleth Maps of the PCA-Based Composite Health-Related Quality of Life Score in Mississippi Counties, 2016 (left panel) and 2025 (right panel) Higher scores (redder shades) indicate worse HRQoL. Maps for 2016 and 2025 are shown to highlight the emergence (2016: Moran’s I = 0.342, p = 0.001) and persistence (2025: I = 0.144, p = 0.011) of significant spatial clustering. The 2015 map is omitted as global clustering was non-significant in that year (I = 0.087, p = 0.073).

Both years reveal a strikingly consistent geographic pattern characterized by a pronounced north-south divide. The Mississippi Delta region (northwest and west-central counties) consistently exhibits the poorest HRQoL, with the most severe outcomes concentrated in counties such as Holmes, Humphreys, Claiborne, Jefferson, Leflore, Coahoma, Quitman, and Tunica. This area forms a clear hotspot of poor health-related quality of life that shows remarkable persistence over the decade.

In contrast, the Gulf Coast and adjacent southern counties (particularly Lamar, Jackson, Hancock, Harrison, Stone, George, and Pearl River) along with suburban counties near Memphis (DeSoto, Madison, Rankin) display the best HRQoL, appearing as distinct cold spots in both periods. Northeastern counties such as Lee and Lafayette also tend toward better outcomes.

Between 2016 and 2025, the overall spatial configuration demonstrates minimal change, with the Delta remaining the core area of poorest HRQoL and the Gulf Coast/suburban south maintaining its relative advantage. Some local fluctuations are evident (e.g., slight worsening in certain Delta counties like Coahoma and Tunica, and improvement in select southern counties), but the broad regional patterning remains highly stable.

These maps illustrate the deeply entrenched and persistent geographic disparities in health-related quality of life across Mississippi, underscoring structural and historical factors that continue to shape HRQoL outcomes over time.

### Local Spatial Clustering of Composite HRQoL

The Local Indicators of Spatial Association (LISA) cluster maps for the PCA-based composite HRQoL score identify statistically significant (p < 0.05) local spatial clusters in Mississippi counties for 2016 and 2025 (Figure 2). In 2016, clustering was more extensive and diverse, with 22 counties in significant clusters. The dominant pattern featured a High-High (HH) hotspot of poor HRQoL in the western Delta region, comprising eight counties (Coahoma, Humphreys, Leflore, Sharkey, Sunflower, Tallahatchie, Washington, Yazoo). A substantial Low-Low (LL) cold spot of better HRQoL included ten counties, primarily along the Gulf Coast and in select central/suburban areas (Forrest, George, Hancock, Harrison, Hinds, Jackson, Marshall, Pearl River, Perry, Stone). Isolated outliers consisted of one High-Low county (Scott) and three Low-High counties (Carroll, Franklin, Warren), with the remaining 60 counties non-significant.

**Figure 2.**
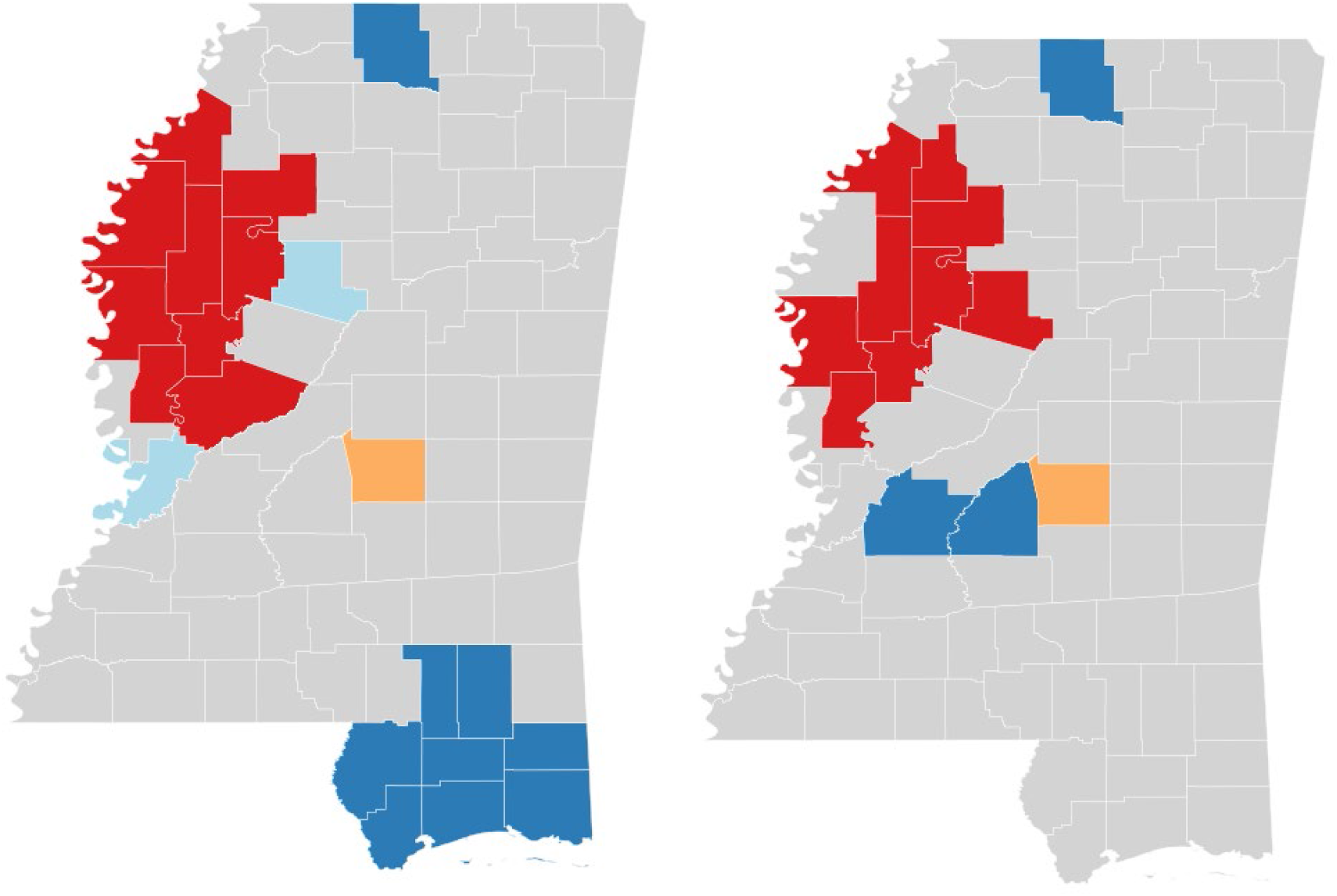
Local Indicators of Spatial Association (LISA) Cluster Maps for the PCA-Based Composite Health-Related Quality of Life Score in Mississippi Counties, 2016 and 2025. Left panel: 2016; Right panel: 2025. Significant clusters (p < 0.05) are shown: High-High (red), Low-Low (blue), High-Low (pink), Low-High (light blue); non-significant counties are gray. Higher composite scores indicate worse HRQoL. Red: High-High (HH); Blue: Low-Low (LL); Pink: High-Low (HL); Light blue: Low-High (LH); Gray: Non-significant

By 2025, clustering intensity had decreased, with only 15 counties in significant clusters. The HH hotspot in the Delta persisted with slight reconfiguration and expansion to nine counties (Carroll, Coahoma, Grenada, Humphreys, Leflore, Quitman, Sunflower, Tallahatchie, Washington), reinforcing the region’s entrenched geographic disadvantage. In contrast, the LL cold spot contracted markedly to just four counties (Hinds, Marshall, Rankin, Stone), with many formerly advantaged Gulf Coast and southern counties reclassified as non-significant. Outliers remained uncommon, including one High-Low (Scott) and one Low-High (Attala) county, leaving 67 counties non-significant.

Overall, these LISA results reveal persistent yet evolving local spatial clustering of HRQoL disparities. The Delta region consistently emerges as a stable High-High hotspot of poor HRQoL across both years, highlighting deep-rooted structural inequities. Meanwhile, the substantial weakening of the Low-Low cold spot along the Gulf Coast and in suburban counties by 2025 suggests erosion of relative advantages in these areas—potentially reflecting statewide diffusion of modest HRQoL improvements (e.g., in physical and mental unhealthy days) or persistent barriers that limited further gains in formerly advantaged regions. The overall reduction in significant clusters from 22 to 15 indicates a slight diffusion of local spatial patterns over the decade, even as core hotspots remain remarkably stable.

### Quantitative Confirmation of Spatial Patterns

Table 3 presents results from a maximum-likelihood spatial lag (SAR) model with year fixed effects, using the PCA-based composite HRQoL score as the dependent variable. The model confirms substantial spatial dependence, with a positive and highly significant spatial lag coefficient (ρ = 0.125, p < 0.001), indicating that a county’s HRQoL is influenced by the HRQoL levels of neighboring counties even after controlling for observed covariates.

**Table 3.** Spatial Autoregressive (SAR) Model Results for the PCA-Based HRQoL Composite Score (2015–2025)

| Variable | Coefficient | Std. Error | z-value | p-value |
| --- | --- | --- | --- | --- |
| Constant | -8.10 | 0.58 | -14.05 | <0.001 |
| Ratio of population to dentists | 0.00 | 0.00 | 1.66 | 0.097 |
| Preventable hospital stays | 0.00 | 0.00 | 2.58 | 0.010 |
| Adult smoking (%) | 29.06 | 1.13 | 25.77 | <0.001 |
| Diabetes prevalence (%) | 3.12 | 1.10 | 2.84 | 0.005 |
| Injury deaths (per 100,000) | 0.00 | 0.00 | 2.56 | 0.010 |
| % Non-Hispanic Black | 0.99 | 0.19 | 5.25 | <0.001 |
| % 65 and older | -1.26 | 1.03 | -1.23 | 0.219 |
| Uninsured adults (%) | 7.68 | 1.19 | 6.44 | <0.001 |
| Median household income | 0.00 | 0.00 | -9.07 | <0.001 |
| Year 2016 | 0.66 | 0.11 | 6.29 | <0.001 |
| Year 2017 | 0.92 | 0.11 | 8.52 | <0.001 |
| Year 2018 | 1.20 | 0.12 | 9.90 | <0.001 |
| Year 2019 | 1.00 | 0.19 | 5.39 | <0.001 |
| Year 2020 | 1.12 | 0.18 | 6.08 | <0.001 |
| Year 2021 | 0.08 | 0.18 | 0.41 | 0.682 |
| Year 2022 | 0.78 | 0.17 | 4.69 | <0.001 |
| Year 2023 | 1.21 | 0.15 | 7.87 | <0.001 |
| Year 2024 | 1.27 | 0.16 | 8.08 | <0.001 |
| Year 2025 | 1.89 | 0.17 | 11.28 | <0.001 |
| $\rho$ (spatial lag) | 0.13 | 0.04 | 3.38 | <0.001 |

Notes: N = 899 county-year observations. Pseudo $R^2 = 0.840$ . Spatial pseudo $R^2 = 0.840$ .
Coefficients represent direct effects. Higher hrqol\_pca values indicate worse health-related quality of life. Year 2015 is the reference category.

Key predictors of worse HRQoL included adult smoking rate (strongest effect), uninsurance rate, diabetes prevalence, % non-Hispanic Black, injury deaths, and preventable hospital stays, while higher median household income was strongly protective. The percentage of residents aged 65 and older and the ratio of population to dentists showed no significant association.

Year fixed effects were largely positive and significant relative to 2015, with the largest increases observed in 2025 (coefficient = 1.893, p < 0.001), suggesting a general worsening of composite HRQoL over time, particularly in the most recent period. The high pseudo R^2^ (0.840) and inclusion of spatial spillovers underscore the model’s strong explanatory power for geographic variation in HRQoL.

## Discussion

This study provides a comprehensive longitudinal examination of health-related quality of life (HRQoL) in Mississippi from 2015 to 2025, using a principal component analysis (PCA)-derived composite index from County Health Rankings & Roadmaps data. Findings reveal a complex trajectory: self-rated poor or fair health remained persistently higher in Mississippi than national averages (stable gap of 0.06–0.07 percentage points), reflecting deep-rooted structural inequities (1, 3, 14, 15). In contrast, recent releases (2023–2025, reflecting approximately 2020–2022 conditions) showed convergence in poor physical unhealthy days and a reversal in mentally unhealthy days, with the latter favoring Mississippi’s most disadvantaged subgroups (e.g., high child poverty, low education, high unemployment).

This reversal may reflect enhanced community resilience, strong social networks, or temporary relief from federal pandemic-era supports (CARES Act, American Rescue Plan), which mitigated acute stressors in socioeconomically vulnerable populations (16).

However, the lack of parallel improvement in self-rated health indicates that short-term symptom relief has not translated into broader perceptions of well-being, likely due to ongoing exposure to poverty, racial inequities, and underinvestment in health infrastructure.

Choropleth maps and Local Indicators of Spatial Association (LISA) confirmed enduring geographic disparities. The Mississippi Delta consistently emerged as a stable High-High hotspot of poor HRQoL across both 2016 and 2025, underscoring entrenched structural disadvantage in this region. In contrast, the Low-Low cold spot of better HRQoL along the Gulf Coast and in suburban counties (e.g., DeSoto, Rankin, Harrison) weakened substantially by 2025, contracting from ten counties in 2016 to just four (Hinds, Marshall, Rankin, Stone), with many formerly advantaged coastal/southern counties reclassified as non-significant. This erosion of relative advantages in the Gulf Coast area suggests diffusion of statewide modest improvements in certain HRQoL domains (particularly symptom-based measures) or persistent barriers that limited further gains in these regions, even as core hotspots in the Delta remained remarkably stable. The spatial autoregressive lag model quantified significant spillover effects (ρ = 0.13, p < 0.001) and identified adult smoking (β = 29.06, p < 0.001) and uninsurance (β = 7.68, p < 0.001) as the strongest modifiable drivers of worse HRQoL, followed by diabetes prevalence and racial composition. A supplementary analysis further demonstrated that counties with comprehensive smoke-free ordinances exhibited markedly lower smoking prevalence and better HRQoL across all indicators. These results align with prior evidence linking tobacco control and insurance coverage to population health gains in the U.S. South (4, 17).

Policy implications are clear and actionable. Expanding comprehensive smoke-free air policies statewide—particularly into the Delta, where such protections are largely absent— represents a high-impact, evidence-based strategy to reduce smoking-related HRQoL burdens. Local ordinances currently protect approximately 37% of the population, but a statewide law remains pending as of early 2026. Similarly, pursuing Medicaid expansion, which Mississippi has not adopted as of January 2026 (with 2025 bills dying in committee amid ongoing legislative discussions), would address the state’s persistently high uninsurance rates and improve access to preventive and chronic disease care, especially in high-burden counties like those in the Delta hotspot. Place-based investments targeting this persistent hotspot (tobacco control, diabetes prevention, primary care enhancement) and sustained support for social determinants of health are essential to convert recent symptom-based gains into lasting improvements in overall well-being, particularly as relative advantages in other regions appear to erode.

### Strengths and Limitations

Strengths of this study include its longitudinal design spanning 11 years of County Health Rankings & Roadmaps data, the construction of a robust PCA-derived composite HRQoL index, the application of advanced spatial analytic techniques (including global Moran’s I, LISA clustering, and spatial autoregressive modeling) to elucidate persistent geographic patterns, and subgroup analyses that reveal nuanced convergence trends across socioeconomic strata.

This study has several limitations, many inherent to the County Health Rankings & Roadmaps ecosystem and Behavioral Risk Factor Surveillance System-derived measures: 2–3 year data lag potentially delaying detection of recent HRQoL shifts; reliance on model-based small-area estimates for key covariates (e.g., adult smoking and uninsurance rates) with varying precision that may attenuate associations during rapid societal changes; county-level aggregation that masks within-county heterogeneity, especially in larger counties such as Hinds and Harrison; and susceptibility of self-reported measures to recall, social desirability, and cultural reporting biases.

Additional study-specific limitations include the PCA-derived composite index assuming linear relationships and being dominated by self-rated poor or fair health, which may underrepresent evolving multidimensional dynamics (e.g., post-pandemic mental health shifts); median imputation of missing covariates risking bias from non-random missingness; dichotomization in subgroup analyses causing information loss and reliance on arbitrary cutpoints; the global spatial autoregressive lag model imposing homogeneous spatial dependence without allowing spatially varying coefficients or multilevel random effects; a predominantly cross-sectional panel structure limiting modeling of true temporal trajectories or lagged effects; and the ecological design precluding individual-level causal inference.

## Conclusion

Despite these limitations, Mississippi’s HRQoL trajectory reflects a tension between recent symptom-based improvements and enduring disparities in overall well-being. The persistent Delta hotspot and erosion of Gulf Coast advantages highlight the need for targeted, place-based interventions to address structural inequities. Policy efforts should prioritize Delta-focused strategies, including expanded tobacco control, diabetes prevention, and insurance coverage to tackle key drivers identified in the SAR model.

Sustained federal and state investments—building on pandemic-era supports—could accelerate convergence, but long-term progress will require confronting deep-rooted inequities to improve not just acute symptoms but holistic perceptions of health in the state’s most vulnerable communities.

## Data Availability

All data are availible from the County Health Rankings & Roadmaps (CHR&R) program website.

https://www.countyhealthrankings.org/explore-health-rankings

## Acknowledgement

The authors declare no potential conflicts of interest with respect to the research, authorship, or publication of this article. The authors received no external financial support for the research, authorship, or publication of this article. No copyrighted material, surveys, instruments, or tools were used in the research described in this article.

## Notes

### Competing Interest Statement

The authors have declared no competing interest.

### Funding Statement

This study did not receive any funding

### Author Declarations

The study used ONLY openly available human data that were originally located at: County Health Rankings & Roadmaps (CHR&R) program website

